# Massive covidization of research citations and the citation elite

**DOI:** 10.1101/2022.01.24.22269775

**Authors:** John P.A. Ioannidis, Eran Bendavid, Maia Salholz-Hillel, Kevin W. Boyack, Jeroen Baas

## Abstract

Massive scientific productivity accompanied the COVID-19 pandemic. We evaluated the citation impact of COVID-19 publications relative to all scientific work published in 2020-2021 and assessed the impact on scientist citation profiles. Using Scopus data until August 1, 2021, COVID-19 items accounted for 4% of papers published, 20% of citations received to papers published in 2020-2021 and >30% of citations received in 36 of the 174 disciplines of science (up to 79.3% in General and Internal Medicine). Across science, 98 of the 100 most-cited papers published in 2020-2021 were related to COVID-19. 110 scientists received >=10,000 citations for COVID-19 work, but none received >=10,000 citations for non-COVID-19 work published in 2020-2021. For many scientists, citations to their COVID-19 work already accounted for more than half of their total career citation count. Overall, these data show a strong covidization of research citations across science with major impact on shaping the citation elite.

## INTRODUCTION

The COVID-19 pandemic resulted in a massive mobilization of researchers across science to address a new major challenge [1]. It is estimated that approximately 4% of the scientific literature published in 2020-2021 was related to COVID-19 [2]: over 720,000 different scientists published over 210,000 relevant publications based on items indexed in Scopus as of August 1, 2021 [2]. COVID-19-related published items exceeded 440,000 by the end of 2021 according to the WHO database [3].

This shift of the research enterprise and massive production of COVID-19-related publications (“covidization”) may have had implications for citations to recent scientific work. In most scientific disciplines, most papers get few, if any, citations in the first year, and citations appear gradually, spread over many years, with citation half-lives that typically exceed 5 years for most scientific fields and may exceed 10 years for some fields [4-6]. The half-life of the citation pattern for COVID-19 work is still unknown, given the short-term follow-up for the COVID-19 published papers. However, the hundreds of thousands of COVID-19 publications likely have drawn citations largely from other very recently published COVID-19 work. Conversely, for non-COVID-19 work, citations from very recent papers (<1-2 years old) are expected to have been a minority. Therefore, it is likely that a large share of citations to very recent work in 2020 and 2021 reflect citations to COVID-19 papers. The extent and distribution of such a COVID-19-enriched pattern of recent citations is worth studying for their implications in understanding the evolving cultural norms. Citations of more recent papers may represent reliance on less vetted, more tentative knowledge. Reliance on less mature knowledge may be more susceptible to reversal, and a number of high-profile retractions have unnerved the scientific world in the COVID-19 era [7].

Moreover, the massive COVID-19 literature and its citations may have had a major impact on the careers of many scientists. The possibility of receiving a large number of citations could be highly appealing to researchers whose careers are influenced by reputation and citation metrics. If covidization of research heralds a new approach to receiving citations, it may change the incentives of scientists motivated by the lure of such scientific rewards. This, in turn, may shift the work of young scientists away from more “gradualist” fields towards COVID-19. The appeal of working on COVID-19, in other words, may extend beyond its health challenges, skewing an important alignment between the burden of disease and interest by scientists.

Here, we compare scientists’ acquisition of citations for COVID-19 and similarly recent non-COVID-19 work; characterize the profiles of scientists that had extraordinary boosts to their citation profiles; and assess whether COVID-19 citations correlated with overall career impact, or whether they had an independent impact in generating a new citation elite. We addressed these questions using comprehensive data from Scopus [8] from 2020-2021.

## RESULTS

### Citations to work published in 2020-2021 and share of citations to COVID-19 work

From January 1, 2020 until August 1, 2021 a total of 5,728,015 items were published and indexed in Scopus, including 210,183 (4%) items related to COVID-19. The number of total citations that they received until August 1, 2021 was 9,174,336, of which 1,832,477 citations (20%) were to the published items related to COVID-19. Therefore, even though COVID-19 items were a minority, they accounted for a 5-times larger share of the citations received to very recently published items.

Table 1 shows the 36 scientific disciplines (of a total of 174 fields across all science) where more than 30% of citations received in 2020-2021 to work published in these two years were to COVID-19 work. For 3 scientific fields, more than two-thirds of the citations received to 2020-2021 were for COVID-19-related work: General & Internal Medicine 79.3%, Virology 76.7%, and Emergency & Critical Care Medicine 66.8%. Stated differently, less than one third of citations in these fields during 2020-2021 referenced non-COVID-19 literature, including literature from all other diseases.

**Table 1.** Scientific disciplines where COVID-19 work received >30% of the citations given to papers published in 2020-2021 (until August 1, 2021)

| Scientific discipline | Items published in 2020-2021 | COVID-19 items published (%) | Citations received to Items published | Citations to COVID-19 items (%) |
| --- | --- | --- | --- | --- |
| General & Internal Medicine | 125,491 | 21,099 (17) | 495,196 | 392,681 (79) |
| Virology | 20,076 | 4,240 (21) | 112,723 | 86,407 (77) |
| Emergency & Critical Care Medicine | 17,921 | 3,155 (18) | 41,399 | 27,654 (67) |
| Tropical Medicine | 13,241 | 1,886 (14) | 35,529 | 23,254 (66) |
| Epidemiology | 6,906 | 1,642 (24) | 20,099 | 12,828 (64) |
| General Clinical Medicine | 11,684 | 1,593 (14) | 20,604 | 13,059 (63) |
| Microbiology | 84,347 | 10,623 (13) | 326,174 | 203,465 (62) |
| Respiratory System | 29,206 | 3,521 (12) | 73,569 | 42,997 (58) |
| Pediatrics | 30,127 | 3,013 (10) | 42,647 | 24,455 (57) |
| Otorhinolaryngology | 18,684 | 1,626 (9) | 22,046 | 12,500 (57) |
| Psychiatry | 42,893 | 4,847 (11) | 97,234 | 52,158 (54) |
| Allergy | 7,474 | 742 (10) | 21,473 | 10,343 (48) |
| Immunology | 56,436 | 5,181 (9) | 208,502 | 96,864 (47) |
| Environmental & Occupational Health | 4,619 | 620 (13) | 5,067 | 2,317 (46) |
| Nuclear Medicine & Medical Imaging | 47,979 | 2,758 (6) | 87,276 | 39,580 (45) |
| Public Health | 46,141 | 6,190 (13) | 63,333 | 27,337 (43) |
| Cardiovascular System & Hematology | 87,112 | 6,786 (8) | 182,546 | 78,478 (43) |
| Dermatology & Venereal Diseases | 29,796 | 2,199 (7) | 34,284 | 14,461 (42) |
| Anesthesiology | 21,658 | 2,433 (11) | 32,644 | 13,577 (42) |
| Geriatrics | 10,825 | 1,139 (11) | 20,734 | 8,205 (40) |
| Arthritis & Rheumatology | 22,188 | 1,595 (7) | 35,697 | 13,790 (39) |
| Medical Informatics | 14,592 | 1,915 (13) | 28,145 | 10,768 (38) |
| Surgery | 51,521 | 3,459 (7) | 58,520 | 21,731 (37) |
| Pathology | 9,479 | 482 (5) | 16,313 | 5,924 (36) |
| Gastroenterology & Hepatology | 38,427 | 2,810 (7) | 81,915 | 29,014 (35) |
| Obstetrics & Reproductive Medicine | 37,973 | 2,270 (6) | 45,199 | 16,002 (35) |
| Applied Ethics | 7,735 | 1,027 (13) | 7,863 | 2,772 (35) |
| Urology & Nephrology | 31,929 | 1,898 (6) | 41,758 | 14,512 (35) |
| Nursing | 48,218 | 3,992 (8) | 28,855 | 9,891 (34) |
| Clinical Psychology | 10,797 | 709 (7) | 16,865 | 5,769 (34) |
| Endocrinology & Metabolism | 36,541 | 2,356 (6) | 87,770 | 29,581 (34) |
| Sport, Leisure & Tourism | 9,808 | 882 (9) | 19,323 | 6,315 (33) |
| Toxicology | 39,493 | 3,163 (8) | 78,274 | 25,359 (32) |
| Substance Abuse | 10,825 | 751 (7) | 16,141 | 5,182 (32) |
| Biophysics | 8,558 | 594 (7) | 20,885 | 6,606 (32) |
| Gender Studies | 3,473 | 215 (6) | 2,159 | 654 (30) |

Across all disciplines, 98 of the 100 most-cited publications published in 2020-2021 were COVID-19-related. Similarly, 97 of the 100 most-cited publications published in 2020 were COVID-19 related. The proportion declined to 76 of the 100 most-cited among publications published in 2021.

As shown in Figure 1, the proportion of papers receiving very high numbers of citations by August 1 of the next calendar year increased very slightly between 2017 and 2019. However papers published in 2020 had a major shift, with much larger proportions of papers receiving very high numbers of citations. The shift was entirely attributable to COVID-19-related publications. Of the 3,183,277 publications in 2020, the 96,351 COVID-19 related publications received 8.4-fold more citations than the non-COVID-19 publications. The fold difference was 20.9-fold for General and Internal Medicine, i.e. on average a COVID-19-related paper received more than 20 times the number of citations received by a non-COVID-19 paper. The average citations per paper were higher for COVID-19 papers than for non-COVID-19 papers for 128 of the 129 scientific disciplines that published more than 50 COVID-19-related papers in 2020 (with the exception of Computational Theory and Mathematics) (Supplementary Table 1).

**Figure 1.**
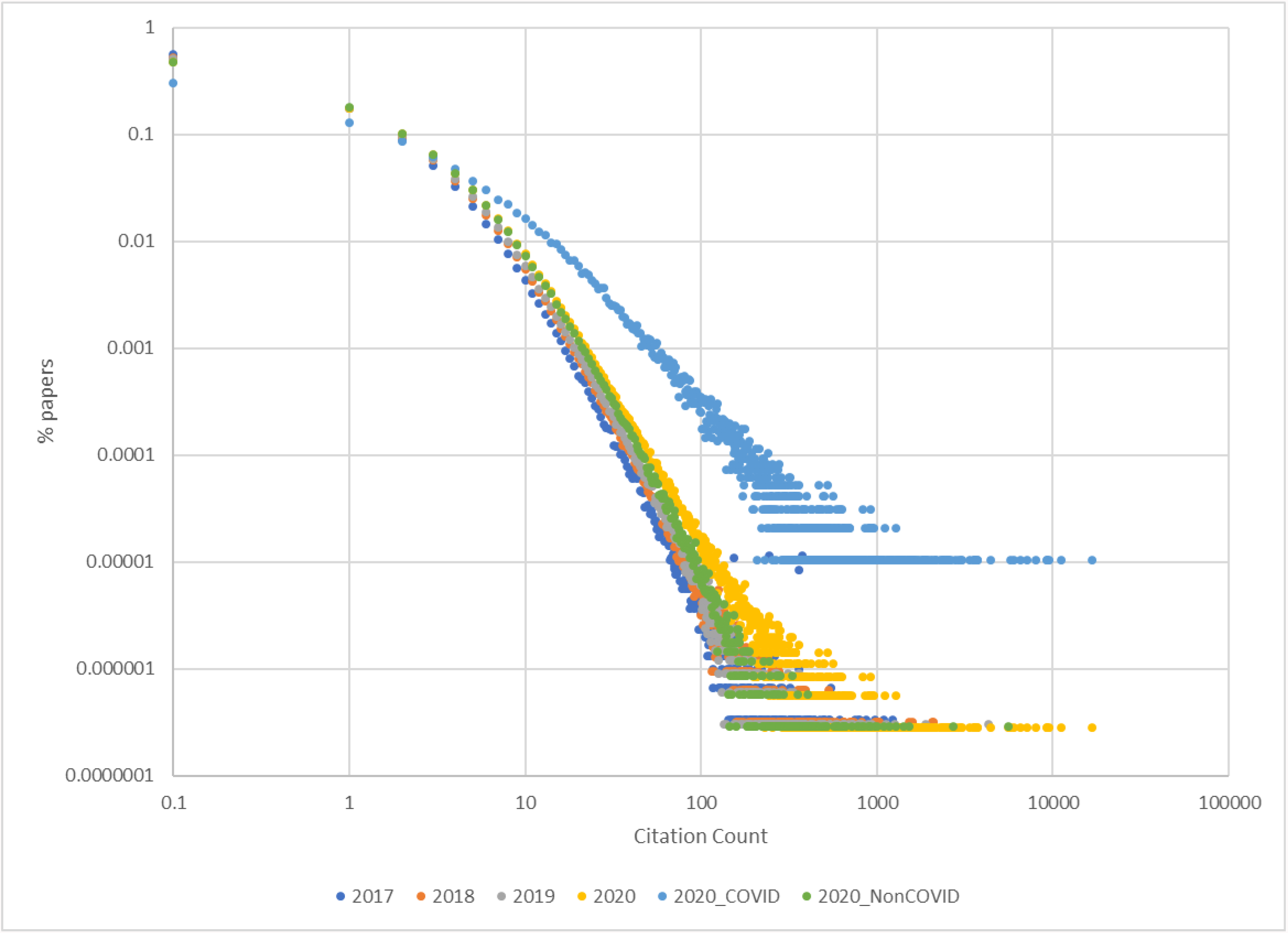
Distribution of the publications with different numbers of citations until August 1 of the next calendar year for publications published in 2017, 2018, 2019, and 2020. For publications published in 2020, separate data are shown for COVID-19 publications and non-COVID-19 publications.

### Scientists with high numbers of citations to their 2020-2021 published work

A total of 84757 scientists had received >=100 citations to their work published in 2020-2021 by August 1, 2021 (among a total of 4,183,909 Scopus IDs that had published at least one paper in that time period and 5 or more papers in their entire career). Among these 84757 scientists, 35358 had received >=300 citations, 5773 had received >=1000 citations, 240 had received >=5000 citations, and 110 had received >=10000 citations for such very recent work.

Of the 84757 scientists with >=100 citations to very recent work, 53% had published at least some COVID-19 papers and of the 5773 scientists with >=1000 citations to very recent work, 65% had published some COVID-19 work. Table 2 shows the number of scientists who had received high numbers of citations to very recent work overall, COVID-19-related work, and non-COVID-19 work. As shown, the number of authors who received >=100 citations to very recent work was almost double for COVID-19 work than for non-COVID-19 work, but the difference was eliminated at the >=1000 citations threshold. >=5000 citations were received only for COVID-19 work, with the exceptions of 6 scientists (three authors of an annual cancer statistics reference, and three authors of cardiology guidelines). At the >=10,000 citation threshold, all 110 scientists conducted COVID-19 work; only 15 of these scientists had also received >=100 citations for their very recent non-COVID-19 work.

**Table 2.** Number of scientists who received high numbers of citations to COVID-19 work and to non-COVID-19 work published in 2020-2021 (data from Scopus until August 1, 2021)

|  | All published work<br>in 2020-2021 | COVID-19 work<br>in 2020-2021 | Non-COVID-19 work<br>in 2020-2021 |
| --- | --- | --- | --- |
| $\geq 100$ citations | 84757 | 30307 | 54891 |
| $\geq 1000$ citations | 5773 | 2923 | 2558 |
| $\geq 5000$ citations | 240 | 229 | 6 |
| $\geq 10000$ citations | 110 | 110 | 0 |

Almost all of the scientists who had received >10,000 citations to their very recent work were from China. In detail, their main affiliation was in China for 104 scientists and Hong Kong for another 4. The 8 most-cited papers of 2020-2021 were all papers from China published in the early days of the pandemic and describing clinical characteristics and preliminary epidemiological features of COVID-19.

### Boosting of career citation impact by citations to COVID-19 work

Among the 84757 scientists with >=100 citations to their very recent work (published in 2020-2021), for n=11767 scientists the citations to their COVID-19 work already accounted for more than half of their total career citation count. Correspondingly, for n=5071 of the 84757 scientists the citations to their non-COVID-19 work published in 2020-2021 already accounted for more than half of their total career citation count.

### Improved overall citation ranking for scientists with influential COVID-19 work

Using a composite citation indicator for ranking the citation impact of scientists, among the top-300 ranked scientists for their COVID-19 work, 117 were among the top-100,000 ranked science-wide for their entire career impact as of August 1, 2021 and 54 were among the top-20,000 ranked science-wide for their career impact. Figure 2 shows the trajectory for the ranking of these 54 scientists across science according to the composite indicator considering the citations received in a single year to all work published in their past career. As shown, in 2019 versus 2017 improvements in ranking were as common as worsening ranking: 13/54 scientists improved their ranking by a third or more and 10/54 worsened their ranking by as much. Conversely, in the 2020 versus 2019 comparison, 47/54 scientists improved their ranking by a third or more, while no scientists worsened their ranking by this margin. Five scientists improved their ranking more than 6-fold; the most impressive improvement was for a long-time coronavirus expert who went from rank 48045 in 2019 to rank 362 in 2020, a 13-fold improvement.

**Figure 2.**
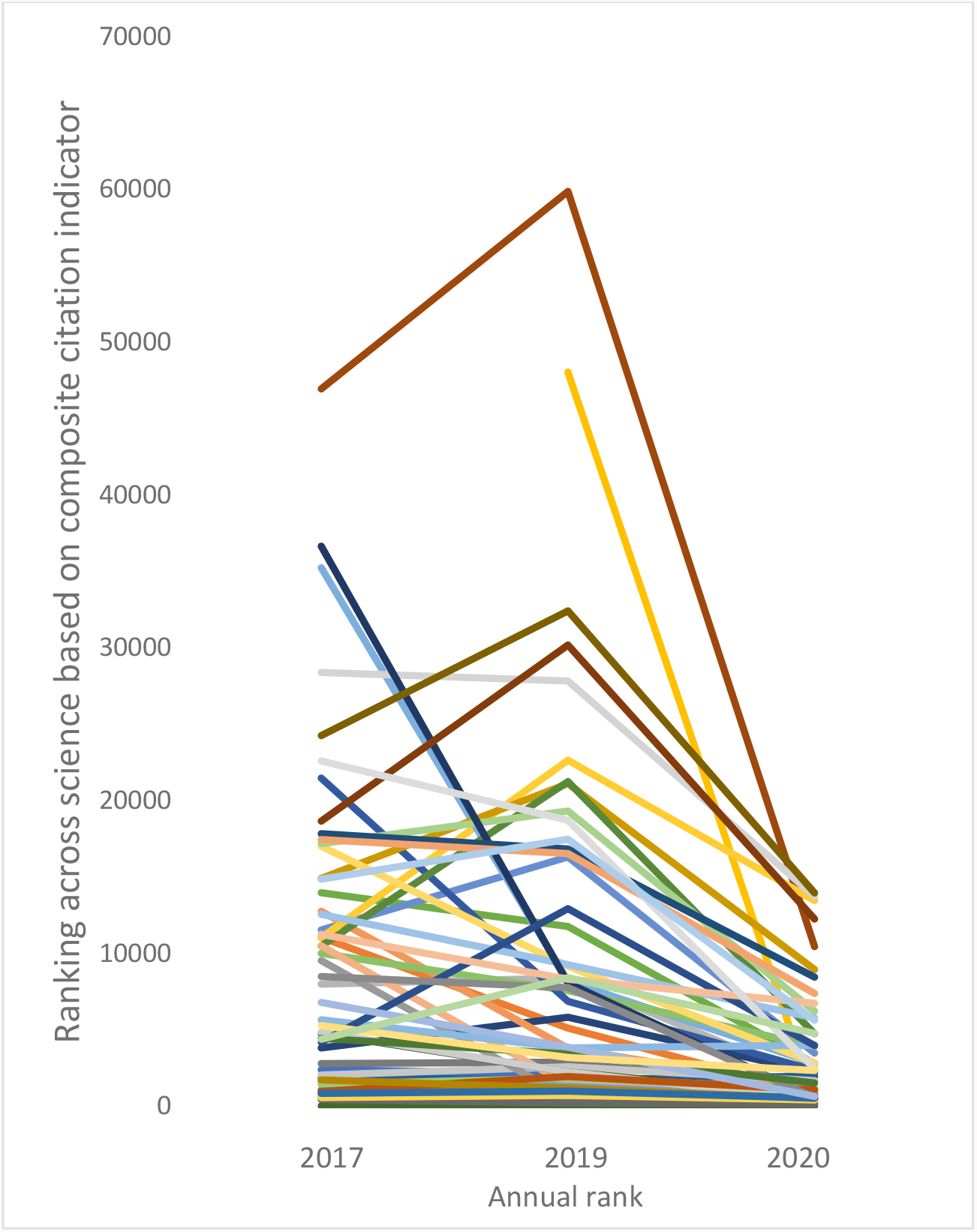
Trajectory of annual ranking (based on composite citation indicator) for 54 scientists who were among the top-300 for the citation impact of their COVID-19 publications and among these top-20,000 for the cumulative citation impact of their work by August 1, 2021.

Overall, 143 of the top-200 ranked scientists across science based the composite citation indicator calculated specifically for their work published in 2020-2021 had published at least 1 COVID-19-related paper.

### Correlation between metrics of impact: career impact and 2020-2021 work

Across the 84757 scientists, the performance for the recent work in 2020-2021 correlated strongly with the respective performance during their entire career, for the number of papers (r=0.71), the ranking according to the composite citation indicator (r=0.57), the h-index (r=0.56), and the hm-index (r=0.59), but not for the number of citations (r=0.16). The number of citations to recent non-COVID-19 work correlated strongly with the number of citations for the entire career (r=0.46), but the number of citations to very recent COVID-19 work did not correlate with the number of citations to the entire career (r=-0.03). The number of citations to recent COVID-19 work was slightly negatively correlated with the number of citations to recent non-COVID-19 work in 2020-2021 (r=-0.14).

The lack of correlation between performance metrics on COVID-19 work and performance for the entire career was seen also for all other metrics besides citations. Figure 3 shows the strong relationship (r=0.59) between the hm-index for entire career and the hm-index for very recent work overall, but weak relationship (r=0.11) between the hm-index for entire career and the hm-index for COVID-19 work.

**Figure 3.**
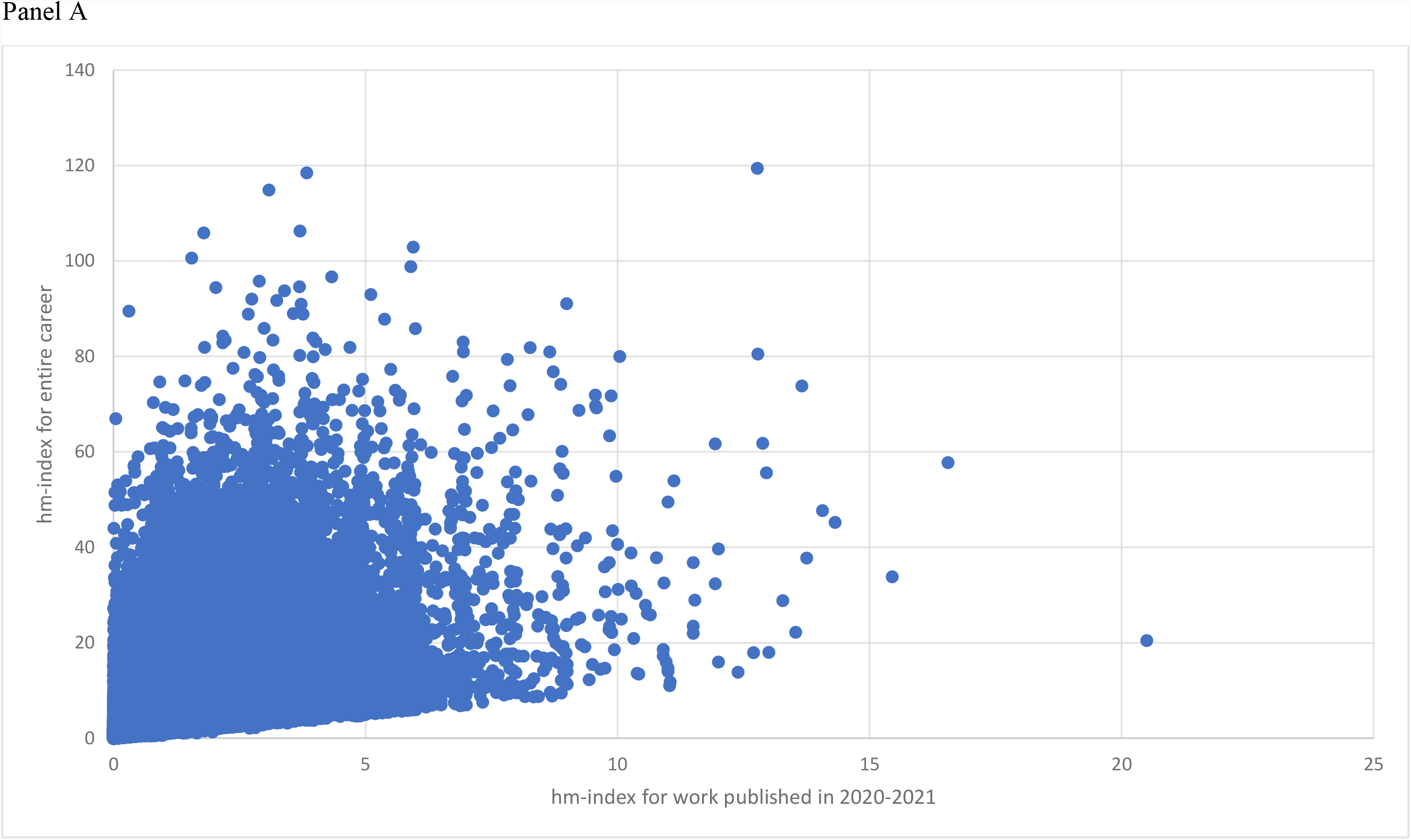
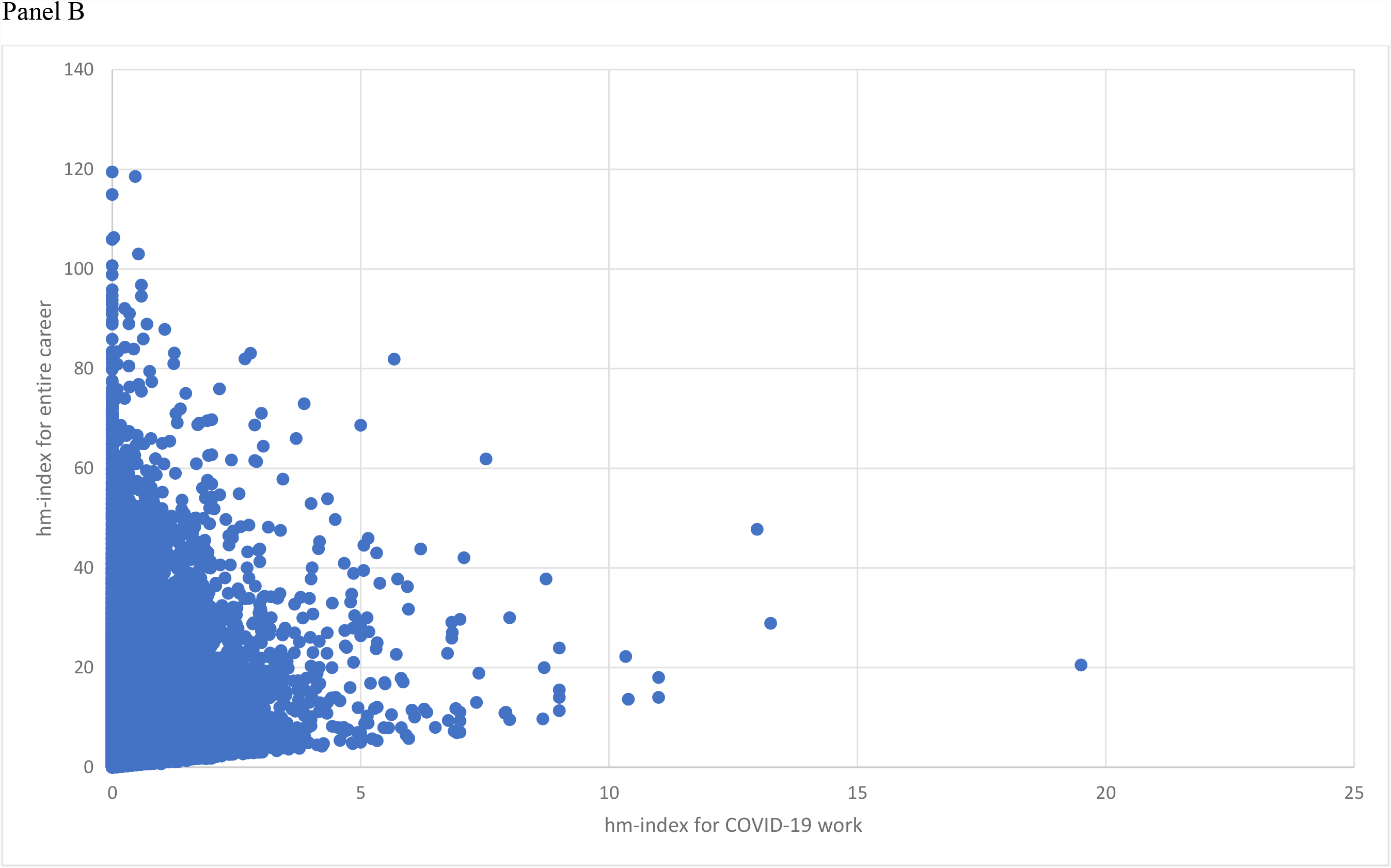
A. Correlation between co-authorship-adjusted hm-index for work published in 2020-2021 and co-authorship-adjusted hm-index for entire career. B. Lack of correlation between co-authorship-adjusted -index for COVID-19 work published in 2020-221 and co-authorship-adjusted hm-index for entire career.

## DISCUSSION

The present analysis shows a massive covidization of research citations during 2020-2021. A large share of the citations to papers published in 2020-2021 has gone to COVID-19-related items. This pattern is seen across many scientific disciplines, with the highest rates in General and Internal Medicine, where >79% of the citations to recent work are to COVID-19 papers. COVID-19 papers published in 2020 received on average more than 8-fold the number of citations than non-COVID-19 papers and the difference exceeded 20-fold in General and Internal Medicine. Almost all of the top-100 most-cited publications published in 2020 across all science (not just biomedicine) were related to COVID-19, and the same applied to three quarters of the most-cited publications published in 2021. Many scientists received in a limited time high numbers of citations to their COVID-19 work and already have higher citations counts for COVID-19 alone than for all other scientific topics combined. The fact that COVID-19 citation impact does not correlate with career-long citation impact means that many authors who are highly-cited for their COVID-19 work have had limited citation impact before the pandemic. COVID-19 is generating a new citation elite and reshaping the existing citation elite of top-cited scientists.

COVID-19 attracted the efforts of both established scientists with strong prior publication and citation record and of new entrants to the scientific literature and young investigators. It is unknown whether these scientists will continue to be heavily involved in COVID-19-related research and whether the vast influence and dominance of COVID-19 on research citations will continue in subsequent years. Perhaps as the pandemic dissipates, the large share of citations to recent work for COVID-19 papers may also dissipate in parallel. The appearance of far more non-COVID-19 related publications among the list of the most-cited papers in 2021 as opposed to the respective list of 2020 is an early indication of such a decline. However, even among the 2021 cohort, still three-quarters of the most-cited publications were COVID-19-related. With COVID-19 papers attracting on average 8 times more citations than other papers in the short-term (up to 20 times more in General and Internal Medicine) and with a given that journals and editors are anxious to boost their impact factors, the attraction for publishing COVID-9 work must have been very strong. The rate of future dissipation of COVID-19 dominance in the top-cited scientific literature is unknown. It is possible that major emphasis on COVID-19 may continue to run strong, regardless of the levels of epidemic activity. Re-shuffling of research resources and the commitment of large amounts of funding to COVID-19 work and related fields may induce a more lasting impact on the scientific literature and its citation footprint.

Citation impact may not necessarily mean high quality or validity of the cited work. Many empirical evaluations of quality aspects of different segments of the COVID-19 scientific literature have consistently shown low quality [9-18]. To our knowledge, there is no large-scale assessment of the correlation between quality scores (with all the difficulty of obtaining such scores) and citation impact of COVID-19 work specifically. However, other investigators have found that COVID-19 papers published in the most influential journals have weaker designs than non-COVID-19 papers in the same venues [15]. Moreover, several extremely-cited COVID-19 papers reflect topics that are debated or even refuted, such as editorials about the origin of the new coronavirus and early reports claiming effectiveness for interventions such as hydroxychloroquine that were not subsequently validated for major outcomes (e.g. mortality). High rates of non-replication and refutation for many of the most highly-cited papers have also been presented in the pre-COVID-19 scientific literature [19,20]. Beyond COVID-19 there is debate in the literature in other fields on the extent to which citations are influenced by quality [21-24] and the relative contribution of rigor and relevance is attracting citations [25-27].

There are several limitations to our work. First, the classification into COVID-19 versus non-COVID-19 work may not be perfect. However, it is unlikely that the existence of a border zone of difficult to classify papers and of papers misclassified by our search algorithm would change the big picture of the results. Second, some scientists may have their publications split into two or more Scopus ID files and some Scopus ID files may include papers by more than one author. Nevertheless, Scopus data have high precision and recall (98.1% and 94.4%, respectively) [4], therefore this is unlikely to be a source of major error. Third, we could not evaluate whether the massive advent of the COVID-19 literature and of its citation footprint affected (negatively) the non-COVID-19 literature and its citations. This is very difficult to evaluate since there are dynamic changes in the volume of the Scopus-indexed literature over time, irrespective of COVID-19. Some of these changes are genuine (e.g. due to emergence of some new hot research areas) and others are artifacts of indexing (e.g. more journals are indexed in Scopus over time).

By design, we opted to focus on authors with at least 5 full papers, conference papers or reviews under their belt by August 21, 2021. This choice has been adopted also in previous work [2]. This design probably excluded from the evaluation a substantial number of early career scientists who have not published that many papers yet, but who may have already co-authored COVID-19 work that gathered many citations. Therefore, the number of authors who more than doubled their total career citations would be probably much larger than our estimates, if these authors with few papers were to be added. However, many Scopus author ID files with few items are fragments that belong to larger profiles and considering these ID files would have added spurious noise to the analysis. Moreover, many of the author ID files with few papers may represent people who have an auxiliary role in the research process rather than being key investigators.

Allowing for these caveats, our analysis shows a massive covidization of research citations. Citations are a main coinage used for choices reflecting funding and career advancement in both academia and the wider scientific community and are widely deemed highly desirable [28,29]. Other investigators have expressed concerns about the covidization of research [30,31]. The duration and evolution of the phenomenon are unknown, but they warrant careful monitoring. The ultrafast generation of the broad COVID-19 research community is most welcome to the extent that it serves the needs of scientific investigation and its translation to useful medical and public health interventions and policy. Conversely, if that community grows disproportionally large and/or it remains pervasive even when the pandemic dissipates, challenges may arise.

Evidence from evaluations of citation patterns in very large scientific fields [32] suggests that when scientific fields grow very large, the list of most-cited papers ossifies to become a canon that slows disruption and real progress. COVID-19 offers a unique example of a scientific field that grew to extremely large dimensions extremely fast. The pandemic has shown the great ability of the scientific workforce to shift attention to an acute problem. It is unknown if this versatility can translate also reversely to shifting away from that problem, when it is no longer an acute and major threat – and/or refocusing on different priorities, if such priorities arise. Regardless, given the strong value that publications and citations have on scientists’ funding and career prospects, COVID-19 may continue to have a dominant presence in the scientific literature and its citations well beyond the end of the pandemic.

## METHODS

### COVID-19 and non-COVID-19 work published in 2020-2021

We used a copy of the Scopus database [4] extracted on August 1, 2021. We identified all publications published and indexed in 2020 and 2021 as of that date. We then separated COVID-19 publications from all publications. Similar to previous work [2], COVID-19-related publications were retrieved by searching in articles published in 2019 or later for any of the following terms in the title, abstract, or key words: ‘sars-cov-2’ OR ‘coronavirus 2’ OR ‘corona virus 2’ OR covid-19 OR {novel coronavirus} OR {novel corona virus} OR ‘2019-ncov’ OR ‘covid’ OR ‘covid19’ OR ‘ncovid-19’ OR ‘coronavirus disease 2019’ OR ‘corona virus disease 2019’ OR ‘corona-19’ OR ‘SARS-nCoV’ OR ‘ncov-2019’, with further filtering through the Elsevier International Center for the Study of Research Lab infrastructure to limit to publications indexed (loaded) in Scopus in 2020 or 2021 only, and with the publication year of 2020 or later.

### Citations to work published in 2020-2021

Scopus citations to all publications in 2020-2021, to COVID-19 publications in 2020-2021 and to non-COVID-19 publications in 2020-2021 were counted as of August 1, 2021. As many publications related to COVID-19 have been first disseminated using pre-print servers, we also included preprint publications from ArXiv, SSRN, BioRxiv, ChemRxiv and medRxiv [2]. Citations from or to preprints were not included in any of the counts.

Publications in 2020-2021 were assigned to a discipline based on their journal of publication and according to the Science Metrix classification of science, which is a standard mapping of all science into 21 main fields and 174 subfield disciplines [33,34]. For each of the 174 subfields, we estimated the share of citations received in 2020-2021 by COVID-19 publications published in 2020-2021 against all publications in the same time frame.

We also examined specifically the top-100 most-cited publications published in 2020-2021, in 2020 alone, and in 2021 to identify how many of them were COVID-19-related. We used citation counts in Scopus as of August 1, 2021.

We also generated plots of the proportion of publications receiving different numbers of citations for papers published in 2017, 2018, 2019, and 2020 considering citations received until August 1 of the following calendar year. For 2020, we considered separately COVID-19-related and non-COVID-19 publications. For each of the scientific fields that had published at least 50 COVID-19 publications in 2020, we assessed the ratio of mean citations to COVID-19 publications over mean citations to non-COVID-19 publications. This analysis is slightly biased in favor of non-COVID-19 publications, since very few COVID-19 publications appeared in the first two months of 2020 and thus non-COVID-19 publications had slightly more time available to be cited on average.

### Authors with at least 100 citations to their 2020-2021 published work

We identified all authors who had received by August 1, 2021 at least 100 citations to their work published in 2020-2021. We noted how many of them had published at least 1 COVID-19-related publication. We also noted how many authors had received by August 1, 2021 at least 100 citations to their COVID-19 versus non-COVID-19 work published in 2020-2021. Numbers of authors passing higher citation thresholds (>=500, >=1000, >=5000, >=10000) for these categories were also noted. We examined the country of the authors at the highest citation levels. For all analyses of authors, similar to prior work [2] we only considered those that have published at least 5 papers (articles, conference papers, or reviews) in their career. This allows the exclusion of authors with limited presence in the scientific literature and of author IDs that may represent split fragments of the publication record of some more prolific authors.

### Citation metrics for the 2020-2021 work and for career-long impact

For each author with >=100 citations to their work published in 2020-2021, we also recorded the number of published items in 2020-2021 (publications including preprints, citations to preprints are not recorded), and additional citation indices limited to the impact of the 2020-2021-published work, the Hirsch h-index [35], the co-authorship adjusted Schreiber hm-index [36]. Using a previously published and validated composite citation indicator [37-39] that combines total citations, h-index, hm-index, and three indicators of citations to works as single, single/first, single/first, single/first/last author, we generated a ranking of scientists based on their 2020-2021 work alone. We did the same calculations and generated the respective and rankings limited to COVID-19 work published in 2020-2021.

For each author with >=100 citations to their work published in 2020-2021, we also calculated the same citation metrics and overall ranking across all science as of August 1, 2021 for the work published during their entire career [40]. We evaluated for how many authors their COVID-19 work accounted for at least half of the citations they had received in their entire career; and for how many authors their non-COVID-19 work published in 2020-2021 accounted for more than half of the citations they had received in their entire career.

We had previously generated [2] a list of the top-300 ranked scientists for their COVID-19 work based on the composite citation indicator. We investigated how many of those were also among the top-100,000 ranked science-wide for their entire career impact as of August 1, 2021 and how many were among the top-20,000 ranked science-wide for their career impact. For the scientists who were among the top-20,000 ranked science-wide for their career impact, we also noted their science-wide ranking for their annual citation impact in the single years 2017, 2019, and 2020; data were extracted from previously published, publicly available datasets that use the composite citation indicator for the ranking [37-39]. This allowed to assess the evolution of the trajectory of the ranking of these scientists before the pandemic and during the pandemic. The annual assessments consider all the citations received in a single year to all work published in the scientist’s career. Therefore, they reflect the recent attention not only to the recent work, but also to all past work.

Finally, we calculated Pearson correlation coefficients for the productivity and citation metrics of the scientists for their entire career as of August 1, 2021 and the respective metrics for COVID-19 work, and non-COVID-19 work published in 2020-2021. This allowed to evaluate whether the career impact tracked with their recent COVID-19 work, non-COVID-19 work, or both.

All calculations throughout the paper include self-citations. No statistical tests were used and no p-values are reported, since analyses are descriptive.

## Data Availability

All data produced in the present study are available upon reasonable request to the authors

---

**Supplementary Table 1.** Mean number of citations until August 1, 2021 to publications published in 2020, separately shown for COVID-19 and non-COVID-19 publications. Scientific disciplines are shown in order of decreasing number of COVID-19 publications and only the 129 of the 174 scientific disciplines that had more than 50 COVID-19 publications in 2020 are shown.

| Scientific disciplines | Mean citations for COVID-19 papers | Mean citations for non-COVID-19 papers | Ratio |
| --- | --- | --- | --- |
| General & Internal Medicine | 29.33 | 1.40 | 20.90 |
| Microbiology | 35.34 | 2.51 | 14.07 |
| Cardiovascular System & Hematology | 18.87 | 1.94 | 9.71 |
| Neurology & Neurosurgery | 16.11 | 2.18 | 7.40 |
| Oncology & Carcinogenesis | 12.41 | 2.74 | 4.54 |
| Public Health | 8.66 | 1.35 | 6.39 |
| Psychiatry | 18.89 | 1.76 | 10.76 |
| Immunology | 37.17 | 3.47 | 10.73 |
| Pharmacology & Pharmacy | 7.12 | 1.68 | 4.25 |
| Artificial Intelligence & Image Processing | 2.17 | 1.44 | 1.50 |
| Surgery | 10.45 | 1.16 | 9.00 |
| Nursing | 4.48 | 0.70 | 6.42 |
| Respiratory System | 21.71 | 1.76 | 12.37 |
| Emergency & Critical Care Medicine | 14.54 | 1.40 | 10.36 |
| Education | 3.38 | 0.92 | 3.66 |
| Virology | 47.32 | 2.58 | 18.38 |
| Gastroenterology & Hepatology | 16.60 | 2.17 | 7.64 |
| Pediatrics | 14.54 | 1.01 | 14.38 |
| Anesthesiology | 8.51 | 1.45 | 5.86 |
| Nuclear Medicine & Medical Imaging | 25.69 | 1.68 | 15.32 |
| Dermatology & Venereal Diseases | 10.20 | 1.06 | 9.59 |
| Obstetrics & Reproductive Medicine | 11.11 | 1.24 | 8.93 |
| Endocrinology & Metabolism | 20.64 | 2.61 | 7.91 |
| Toxicology | 17.80 | 2.31 | 7.71 |
| Tropical Medicine | 19.03 | 1.48 | 12.88 |
| Urology & Nephrology | 12.35 | 1.31 | 9.40 |
| Developmental Biology | 49.97 | 4.96 | 10.07 |
| Dentistry | 7.57 | 1.09 | 6.91 |
| Business & Management | 4.89 | 1.45 | 3.36 |
| Epidemiology | 12.80 | 2.00 | 6.40 |
| Otorhinolaryngology | 13.87 | 0.87 | 16.03 |
| Medical Informatics | 12.16 | 2.09 | 5.81 |
| Economics | 7.92 | 1.45 | 5.47 |
| Biochemistry & Molecular Biology | 28.46 | 3.00 | 9.50 |
| Arthritis & Rheumatology | 15.52 | 1.56 | 9.94 |
| Ophthalmology & Optometry | 8.51 | 1.12 | 7.59 |
| General Clinical Medicine | 16.38 | 1.18 | 13.83 |
| Political Science & Public Administration | 4.55 | 0.86 | 5.30 |
| Environmental Sciences | 33.37 | 3.51 | 9.52 |
| Orthopedics | 7.26 | 1.07 | 6.76 |
| Geriatrics | 12.06 | 2.10 | 5.73 |
| Applied Ethics | 3.99 | 0.99 | 4.01 |
| Energy | 8.98 | 2.64 | 3.40 |
| Health Policy & Services | 6.41 | 1.50 | 4.27 |
| Law | 1.25 | 0.49 | 2.54 |
| Networking & Telecommunications | 5.46 | 1.76 | 3.11 |
| Medicinal & Biomolecular Chemistry | 12.09 | 2.19 | 5.51 |
| Strategic, Defence & Security Studies | 6.71 | 2.53 | 2.66 |
| Experimental Psychology | 14.94 | 1.95 | 7.66 |
| Geography | 3.81 | 1.30 | 2.94 |
| Nutrition & Dietetics | 17.17 | 2.17 | 7.93 |
| Rehabilitation | 7.45 | 0.97 | 7.70 |
| Information & Library Sciences | 3.39 | 0.89 | 3.82 |
| Veterinary Sciences | 4.54 | 0.97 | 4.70 |
| Clinical Psychology | 15.52 | 1.61 | 9.62 |
| Bioinformatics | 15.85 | 3.20 | 4.96 |
| Allergy | 27.26 | 2.52 | 10.82 |
| Environmental & Occupational Health | 6.38 | 0.94 | 6.79 |
| Gerontology | 7.28 | 1.80 | 4.04 |
| Social Psychology | 8.97 | 1.41 | 6.34 |
| Biophysics | 13.05 | 2.60 | 5.02 |
| Biotechnology | 10.52 | 3.19 | 3.29 |
| Substance Abuse | 16.34 | 1.68 | 9.75 |
| Plant Biology & Botany | 7.58 | 1.91 | 3.98 |
| Sport, Leisure & Tourism | 17.43 | 2.19 | 7.97 |
| Biomedical Engineering | 11.98 | 2.83 | 4.23 |
| Ecology | 6.04 | 2.12 | 2.85 |
| Analytical Chemistry | 22.20 | 2.76 | 8.05 |
| Legal & Forensic Medicine | 4.21 | 0.44 | 9.67 |
| Computation Theory & Mathematics | 0.64 | 0.81 | 0.79 |
| Organic Chemistry | 14.09 | 3.96 | 3.56 |
| Sport Sciences | 8.96 | 1.65 | 5.44 |
| Nanoscience & Nanotechnology | 30.81 | 6.65 | 4.63 |
| Meteorology & Atmospheric Sciences | 19.29 | 3.07 | 6.29 |
| Pathology | 23.97 | 1.84 | 13.05 |
| Criminology | 5.13 | 0.89 | 5.77 |
| Chemical Physics | 13.36 | 3.34 | 4.00 |
| Applied Mathematics | 7.58 | 2.52 | 3.01 |
| International Relations | 2.16 | 0.73 | 2.95 |
| Materials | 3.23 | 1.98 | 1.63 |
| Developmental & Child Psychology | 8.14 | 1.55 | 5.25 |
| Communication & Media Studies | 3.48 | 1.02 | 3.41 |
| Mathematical Physics | 28.83 | 3.19 | 9.05 |
| Complementary & Alternative Medicine | 8.14 | 1.10 | 7.40 |
| Urban & Regional Planning | 4.89 | 1.56 | 3.13 |
| Sociology | 4.52 | 0.80 | 5.67 |
| Anthropology | 0.81 | 0.69 | 1.19 |
| Social Work | 3.46 | 0.86 | 4.04 |
| Nuclear & Particle Physics | 3.99 | 2.68 | 1.49 |
| Fluids & Plasmas | 15.58 | 2.84 | 5.48 |
| Operations Research | 5.80 | 1.80 | 3.21 |
| Logistics & Transportation | 18.64 | 2.06 | 9.04 |
| Industrial Engineering & Automation | 8.91 | 1.39 | 6.42 |
| Religions & Theology | 1.10 | 0.22 | 5.07 |
| Geological & Geomatics Engineering | 4.79 | 2.04 | 2.35 |
| Languages & Linguistics | 2.47 | 0.49 | 5.08 |
| Food Science | 12.73 | 3.01 | 4.23 |
| Information Systems | 10.58 | 2.13 | 4.96 |
| Finance | 36.54 | 2.27 | 16.12 |
| Marketing | 18.16 | 2.41 | 7.53 |
| Development Studies | 5.93 | 1.32 | 4.50 |
| Statistics & Probability | 7.27 | 1.51 | 4.83 |
| Applied Physics | 3.18 | 2.40 | 1.32 |
| Genetics & Heredity | 10.77 | 2.02 | 5.33 |
| Acoustics | 9.35 | 2.03 | 4.61 |
| Gender Studies | 3.66 | 0.65 | 5.68 |
| Agricultural Economics & Policy | 16.33 | 1.88 | 8.70 |
| Mechanical Engineering & Transports | 5.20 | 1.16 | 4.47 |
| Literary Studies | 0.30 | 0.09 | 3.41 |
| Evolutionary Biology | 16.26 | 2.21 | 7.35 |
| Science Studies | 6.38 | 2.18 | 2.92 |
| Environmental Engineering | 9.91 | 2.31 | 4.30 |
| Physiology | 11.84 | 2.19 | 5.41 |
| General Physics | 6.76 | 2.89 | 2.34 |
| Optics | 6.86 | 2.05 | 3.34 |
| History | 2.73 | 0.19 | 14.66 |
| Human Factors | 10.31 | 2.06 | 5.01 |
| Building & Construction | 5.20 | 2.42 | 2.15 |
| General Mathematics | 3.51 | 1.16 | 3.03 |
| Psychoanalysis | 0.75 | 0.28 | 2.68 |
| Polymers | 5.16 | 3.01 | 1.71 |
| Optoelectronics & Photonics | 2.90 | 1.13 | 2.56 |
| Electrical & Electronic Engineering | 1.72 | 1.36 | 1.26 |
| Cultural Studies | 1.53 | 0.28 | 5.43 |
| Speech-Language Pathology & Audiology | 4.25 | 1.39 | 3.07 |
| Philosophy | 0.30 | 0.28 | 1.06 |
| Social Sciences Methods | 3.51 | 1.09 | 3.23 |
| Chemical Engineering | 6.46 | 3.40 | 1.90 |
| Family Studies | 6.71 | 1.25 | 5.35 |

## REFERENCES

1. Else H. How a torrent of COVID science changed research publishing - in seven charts. Nature 588, 553.

2. Ioannidis JPA, Salholz-Hillel M, Boyack KW, Baas J. 2021 The rapid, massive growth of COVID-19 authors in the scientific literature. Royal Soc. Open Sci. https://doi.org/10.1098/rsos.210389

3. https://search.bvsalud.org/global-literature-on-novel-coronavirus-2019-ncov/, last accessed December 25, 2021

4. Libkind AN, Markusova VA, Libkind IA. Approach for using Journal Citation Reports in determining the dynamics of half-life indicators of journals, Autom Doc Math Linguist 2020;54:174–183.

5. Gilyarevskii RS, Libkind AN, Libkind IA., et al. The obsolescence of cited and citing journals: half-lives and their connection to other bibliometric indicators. Autom Doc Math Linguist 2021;55:152–165.

6. Davis PM, Cochran A. Cited half-life of the journal literature. arXiv 2015; 1504.07479 [cs.DL].

7. Frampton G, Woods L, Scott DA. Inconsistent and incomplete retraction of published research: A cross-sectional study on Covid-19 retractions and recommendations to mitigate risks for research, policy and practice. PLoS One. 2021;16(10):e0258935.

8. Baas J, Schotten M, Plume A, Côté G, Karimi R. 2020 Scopus as a curated, high-quality bibliometric data source for academic research in quantitative science studies. Quant. Sci. Stud. 1, 377–386. (doi:10.1162/qss_a_00019)

9. Abbott R, Bethel A, Rogers M, Whear R, Orr N, Shaw L, Stein K, Thompson Coon J. 2021 Characteristics, quality and volume of the first 5 months of the COVID-19 evidence synthesis infodemic: a meta-research study. BMJ Evid. Based Med.bmjebm-2021-111710. (doi:10.1136/bmjebm-2021-111710)

10. Bagdasarian N, Cross GB, Fisher D. 2020 Rapid publications risk the integrity of science in the era of COVID-19. BMC Med. 18, 192. (doi:10.1186/s12916-020-01650-6)

11. Balaphas A, Gkoufa K, Daly MJ, de Valence T. 2020 Flattening the curve of new publications on COVID-19. J. Epidemiol. Community Health 74, 766–767. (doi:10.1136/jech-2020-214617)

12. Khatter A, Naughton M, Dambha-Miller H, Redmond P. 2021 Is rapid scientific publication also high quality? Bibliometric analysis of highly disseminated COVID-19 research papers. Learn Publ. (doi:10.1002/leap.1403)

13. Wang D et al. 2021 Abstracts for reports of randomised trials of COVID-19 interventions had low quality and high spin. J. Clin. Epidemiol. 139, 107–120. (doi:10.1016/j.jclinepi.2021.06.027)

14. Li Y et al. 2021 Reporting and methodological quality of COVID-19 systematic reviews needs to be improved: an evidence mapping. J. Clin. Epidemiol. 135, 17–28. (doi:10.1016/j.jclinepi.2021.02.021)

15. Quinn TJ et al. 2021 Following the science? Comparison of methodological and reporting quality of covid-19 and other research from the first wave of the pandemic. BMC Med. 1, 46. (doi:10.1186/s12916-021-01920-x)

16. Luo X et al. 2021 Consistency of recommendations and methodological quality of guidelines for the diagnosis and treatment of COVID-19. J. Evid. Based Med. 14, 40–55. (doi:10.1111/jebm.12419)

17. Yang S, Li A, Eshaghpour A, Ivanisevic S, Salopek A, Eikelboom J, Crowther M. 2020 Quality of early evidence on the pathogenesis, diagnosis, prognosis and treatment of COVID-19. BMJ Evid. Based Med.bmjebm-2020-111499. (doi:10.1136/bmjebm-2020-111499)

18. Nieto I, Navas JF, Vázquez C. 2020 The quality of research on mental health related to the COVID-19 pandemic: a note of caution after a systematic review. Brain Behav. Immun. Health 7, 100123. (doi:10.1016/j.bbih.2020.100123)

19. Ioannidis JP. 2005 Contradicted and initially stronger effects in highly cited clinical research. JAMA 294, 218–28. doi: 10.1001/jama.294.2.218.

20. Open Science Collaboration 2015 Estimating the reproducibility of psychological science. Science 349, aac4716. doi: 10.1126/science.aac4716.

21. Walter G, Bloch S, Hunt G, Fisher K. 2003 Counting on citations: a flawed way to measure quality. Med. J. Aust. 178, 280–1. doi:10.5694/j.1326-5377.2003.tb05196.x.

22. Buela-Casal G, Zych I. 2010 Analysis of the relationship between the number of citations and the quality evaluated by experts in psychology journals. Psicothema. 22, 270–6.

23. Fabre A. 2016 Same quality but not the same impact: Citations related to publications about celiac disease in JPGN and AJG. J Pediatr Gastroenterol Nutr. 62(4):e38. doi: 10.1097/MPG.0000000000001061.

24. Molléri JS, Petersen K, Mendes E. 2018 Towards understanding the relation between citations and research quality in software engineering studies. Scientometrics. 117, 1453–1478. doi: 10.1007/s11192-018-2907-3.

25. Aksnes DW. 2003 Characteristics of highly cited papers. Res. Eval. 12, 159–170.

26. Aksnes DW. 2006 Citation rates and perceptions of scientific contribution J. Assoc. Inform. Sci. Tech. 57, 169–185.

27. Bornmann L, Daniel H-D. 2008 What do citation counts measure? A review of studies on citing behavior. J. Documentation 64, 45–80.

28. Van Wesel M. 2016 Evaluation by citation: trends in publication behavior, evaluation criteria, and the strive for high impact publications. Sci Eng Ethics. 22, 199–225.

29. Baccini A, De Nicolao G, Petrovich E. 2019 Citation gaming induced by bibliometric evaluation: A country-level comparative analysis. PLoS ONE. 14, e0221212.

30. Pai M. 2020 Covidization of research: what are the risks? Nat. Med. 26, 1159. (doi:10.1038/s41591-020-1015-0)

31. Adam D. 2020 Scientists fear that ‘covidization’ is distorting research. Nature 588, 381–382. (doi:10.1038/d41586-020-03388-w)

32. Chu JSG, Evans JA. 2021 Slowed canonical progress in large fields of science. Proc Natl Acad Sci U S A. 118, e2021636118. doi: 10.1073/pnas.2021636118.

33. Archambault É, Beauchesne OH, Caruso J. 2011 Towards a multilingual, comprehensive and open scientific journal ontology. In Proc. of the 13th Int. Conf. of the Int. Soc. for Scientometrics and Informetrics (ISSI), Durban, South Africa, pp. 66–77.

34. Zhang X, Zhao J, LeCun Y. 2015 Character-level convolutional networks for text classification. Adv. Neural Inf. Processing Syst. 28, 649–657.

35. Hirsch JE. 2005 An index to quantify an individual’s scientific research output. Proc. Natl. Acad. Sci. USA 102, 16569–16572.

36. Schreiber M. 2008 A modification of the h-index: The hm-index accounts for multi-authored manuscripts. J. Informatics 2, 211–216.

37. Ioannidis JP, Klavans R, Boyack KW. 2016 Multiple citation indicators and their composite across scientific disciplines. PLoS Biol. 14, e1002501. (doi:10.1371/journal.pbio.1002501)

38. Ioannidis JPA, Baas J, Klavans R, Boyack KW. 2019 A standardized citation metrics author database annotated for scientific field. PLoS Biol. 17, e3000384. doi: 10.1371/journal.pbio.3000384.

39. Ioannidis JPA, Boyack KW, Baas J. 2020 Updated science-wide author databases of standardized citation indicators. PLoS Biol. 18, e3000918. (doi:10.1371/journal.pbio.3000918)

40. Baas J, Boyack K, Ioannidis JPA. 2021 Data for updated science-wide author databases of standardized citation indicators. Versions 1, 2, and 3 (doi:10.17632/btchxktzyw.1, doi:10.17632/btchxktzyw.2, doi:10.17632/btchxktzyw.3)

